# What is the therapeutic quality of exercise programs in chronic low back pain randomized controlled trials assessed by i-CONTENT tool? A meta research study protocol

**DOI:** 10.1101/2023.06.23.23291759

**Authors:** Ignazio Geraci, Silvia Bargeri, Giacomo Basso, Greta Castellini, Alessandro Chiarotto, Silvia Gianola, Raymond Ostelo, Marco Testa, Tiziano Innocenti

**Author notes:** co-first authors (Ignazio Geraci, Silvia Bargeri). Alphabetic order except for co-first, second and last authors.

## Abstract

**Background:** Exercise therapy is a common intervention recommended for chronic low back pain (cLBP). Although adequate reporting of intervention is crucial to understand and replicate exercise therapy, it does not help clinicians to determine the therapeutic quality. So The international Consensus on Therapeutic Exercise aNd Training (i-CONTENT) tool was developed to assess therapeutic quality of exercise. Therefore, we will assess the therapeutic quality of different exercise interventions by i-CONTENT tool in cLBP RCTs and its inter-rater reliability.

**Methods:** We will perform a meta-research study, starting from Cochrane review publication “Exercise therapy for chronic low back pain”. We will select a random sample of 100 arms with different type of exercises included (i.e. Core Strengthening, General Strengthening, Stretching, Aerobic exercises, Motor Control, Pilates, McKenzie, Qigong, Yoga, Tai Chi). For each included study’s arm, two pairs of independent reviewers will assess the therapeutic quality of exercises applying the i-CONTENT tool. We will calculate the percentage agreement between raters to assess inter-rater reliability.

**Ethics and dissemination:** This study does not require an ethics review as we will not collect personal data. The use of the i-CONTENT tool can help assess the therapeutic quality of studies, reducing the risk of ineffective exercise interventions. The study’s results will be published in peer-reviewed journals and presented at national conferences.

## 1. Introduction

Low back pain is an extremely common symptom that cause activity limitation and participation restriction, with a prevalence in 2017 estimated to be around 577.00 million people^1^. It is the leading global cause of years lived with disability since 1990^1^, becoming a public health concern^2^.

Exercise therapy is a very common intervention, especially recommended for chronic low back pain (cLBP). Several studies^2–5^ have shown that exercise therapy, like motor control exercises, strengthening and endurance exercise, is effective as compared to no treatment and usual care for the treatment of cLBP. However, exercise therapy can be differently prescribed in terms of treatment design (e.g., standard, individualised), dose (duration, frequency, intensity), delivery format (e.g., clinician supervised, group), type (e.g. strengthening, stretching), and combination with other conservative treatments^5^. All of these variables should be clearly and completely reported when describing exercise interventions in randomized controlled trials (RCTs) in order to allow replicability of interventions in clinical practice and research. In recent years, different tools, such as the Consensus on Exercise Reporting Template (CERT)^6^ and the Template for Intervention Description and Replication (TIDieR) checklist^7^ have been developed to improve the reporting of exercise interventions in rehabilitation research to enhance exercise reproducibility and clinical translation. However, these currently available reporting tools do not interpret the therapeutic quality (i.e., ‘the potential effectiveness of a specific intervention given the potential target group of patients’) of exercise interventions. To yield optimal effects, the content of an exercise programme should be in line with the latest research, be tailored to the potential of the participants^8^ and be of sufficient volume ^9,10^. For instance, studies of the dose responsiveness of strength training clearly indicate that strength training programmes produce the greatest increases in muscle strength when the training load is high^10^. So systematic reviews of interventions designed to increase muscle strength should assess whether the training load was adequate. In 2020, The international Consensus on Therapeutic Exercise aNd Training (i-CONTENT) tool^11^ was developed for this purpose to assess by a rating tool, instead of a reporting guideline, the risk of ineffectiveness of the exercise purposed and to better identify, appraise and interpret the heterogeneity across RCTs of exercise.

### 1.2 Objectives

The aim of this study will be to assess the therapeutic quality of exercise interventions with i-CONTENT tool in cLBP RCTs and to assess its inter-rater reliability ^12^.

## 2. Methods Study design

We will perform a meta-research study. Since that the specific reporting checklist is under development^13^, we will adapt items from the Preferred Reporting Items for Systematic reviews and Meta-Analyses (PRISMA) checklist^14^ for the reporting of this study.

### 2.1 Eligibility criteria and information sources

We will start from RCTs included in the 2021 Cochrane review publication “Exercise therapy for chronic low back pain”^5^ to select a random sample of 100 exercise arms of different type of exercises (e.g, core strengthening, general strengthening, flexibility, aerobic exercises, pilates, McKenzie, Qigong, Tai Chi, yoga). In case of mixed interventions where exercise is combined with other conservative treatments (e.g. drugs, electrotherapy), we will exclude the related study’s arm if exercise comprised <75% of the treatment (per judgement of the extractor). To ensure consistency of judgments, mixed exercises type (e.g. aerobic plus core strengthening) will be excluded.

### 2.2 Data management

Two reviewers (IG, SB) will extract the following characteristics: author, year of publication, country, sample size of the arms, population characteristics (e.g., age, sex), symptom duration (e.g., mean months), presence of radicular symptoms/leg pain, intervention (e.g., type of exercises, frequency, intensity), comparison and outcomes assessed.

### 2.3 Application of the i-CONTENT tool

According to the i-CONTENT tool^11^, to yield the potential effectiveness of a therapeutic exercise, the exercise programme should match the patients’ problems, should be based on a proven rationale to determine its optimal frequency, intensity, time and type, should be applied by a qualified supervisor, assessed with a proper outcome measure, being safe, and with an adequate therapy adherence.

For each included study’s arm, two pairs of independent reviewers (IG, GC) (SB, GB) will be involved in the assessment of the therapeutic quality of exercises applying the i-CONTENT tool^11^. The sample will be divided in four subsets (**Table 1**). They will independently assess seven items: (i) patient selection, (ii) dosage of the exercise programme, (iii) type of exercise programme, (iv) qualified supervisor, (v) type and timing of outcome assessment, (vi) safety of the exercise programme and (vii) adherence to the exercise programme.

**Table 1.**

|  | Set A (25%) | Set B (25%) | Set C (25%) | Set D (25%) |
| --- | --- | --- | --- | --- |
| Rater 1 | X |  |  | x |
| Rater 2 | x | X |  |  |
| Rater 3 |  | x | X |  |
| Rater 4 |  |  | x | X |

All items will be evaluated as “low risk of ineffectiveness” or “high risk of ineffectiveness” of the exercise intervention. If no details on the topic will be reported, items will be judges as ‘probably done’ or ‘probably not done’. Each evaluation will be substantiated by a rationale to support the evaluation. A calibration phase will be done on four RCTs on different exercise type. Any disparities were resolved by consensus discussion with another pair of reviewers (TI, SG).

The full checklist along with instructions used by reviewers for judgements is reported in **Appendix A**.

## Data Availability

All data produced in the present work are contained in the manuscript

## Statistical analysis

Data will be presented descriptively in tabular form as tables and figures. We will use descriptive statistics to describe the proportion of items assessed as “low risk” or “high risk” of ineffectiveness, “probably done” or “probably not done” in all exercise arms, as well as for each exercise type. To assess the inter-rater reliability, we will calculate the percentage agreement as well as specific positive and specific negative agreement according to de Vet et al^12^ between two raters. A percentage of agreement ≥70% will be considered satisfactory ^12^.

All data analyses will be performed using STATA.

## Ethics and dissemination

This study does not require an ethics review as we will not collect personal data.

Tools such as the i-CONTENT tool can enable clinicals and researchers to assess the therapeutic quality of studies, to avoid high risk of ineffectiveness when planning an exercise interventions. Results of this study will be published in a peer-reviewed journals and disseminated in national conference presentations. Through this judgment it is possible to establish whether that exercise can be effective and adequate to that patient.

## Author contributions

TI, IG, SB, GB conceived and designed the study protocol.

IG, SB, GB, GC, AC, SG, RO, TI were involved in conceptualising the study objectives, providing input into study selection criteria and plans for data extraction. All the authors, including SB, GB, IG, GC, AC, SG, RO, MT, TI revised and approved the final version of the protocol.

## Funding statement

This research received no specific grant from any funding agency in the public, commercial or notfor-profit sectors. SB, SG, GC were supported by the Italian Ministry of Health. The funding source had no controlling role in the study design, data collection, analysis, interpretation, or report writing

## Competing interest

None

## Appendix A. Checklist i-CONTENT tool

|  | **“Low risk” of ineffectiveness** | **“High risk” of ineffectiveness** | If not details<br>Probably<br>(done) | on the topic<br>Probably not<br>(done) | Support for judgement |
| --- | --- | --- | --- | --- | --- |
| **Patient selection** | ☐ The purpose of the exercise therapy program matches the patients’ problems (directly or through a plausible causative relationship). | ☐ The purpose of the exercise therapy program does not match the patients’ problems. | ☐ | ☐ |  |
| **Dosage of the Exercise program** | ☐ The investigators applied a plausible or proven rationale* to determine the ‘Frequency’, ‘Intensity’, and ‘Time’ of the exercise program, matching the purpose of the exercise intervention. | ☐ The investigators did not use a plausible or proven rationale*, did not match the rationale with the purpose of the exercise program, or did not match the rationale and the ‘Frequency’, ‘Intensity’, and ‘Time’ of the exercise program. | ☐ | ☐ |  |
| **Type of the Exercise program** | ☐ The investigators applied a plausible or proven rationale* to determine the ‘Type’ of exercise, defined as the form in which the exercise is provided, and the investigators matched the ‘Type’ of the exercise therapy program with the purpose of the exercise therapy program. | ☐ The investigators did not use a plausible or proven rationale* or did not match the ‘Type’ of the exercise program with the purpose of the exercise therapy program. | ☐ | ☐ |  |
| **Qualified Supervisor (if applicable)** | ☐ The supervisors of the exercise therapy program are experienced with the targeted patient population and sufficiently skilled in providing the proposed exercise program. | ☐ The supervisors of the exercise therapy program are inexperienced with the patient population or insufficiently skilled to provide the exercise program | ☐ | ☐ |  |
| **Type and Timing of Outcome Assessment** | ☐ The investigators used one or more valid and responsive performance-based outcome measure(s) which reflect the goals and purpose of the exercise program to assess the effectiveness exercise therapy program. The measurements have taken place within the time window where the expected effect would most likely take place. | ☐ The investigators use a non- validated performance measure as primary outcome measure to assess the effect of the therapeutic intervention. | ☐ | ☐ |  |
| **Safety of the Exercise Program** | ☐ The number and severity of the exercise- related adverse events in the study are in line with the expected number of adverse events for similar exercise programs in similar populations. | ☐ The number and severity of the exercise related adverse events are substantially higher than what would be expected. | ☐ | ☐ |  |
| **Adherence to the Exercise Program** | ☐ The intended exercise dosing was achieved, based on relevant information regarding to exercise adherence (i.e., the number of sessions attended, the number of exercises performed, and whether or not the intended exercise dosage was reached). | ☐ The level of exercise adherence of patients to the exercise therapy program was insufficient to assume the intended exercise dosing was achieved. | ☐ | ☐ |  |

## Notes

### Competing Interest Statement

The authors have declared no competing interest.

